# Impact of Nasopharyngeal Specimen Quality on SARS-CoV-2 Test Sensitivity

**DOI:** 10.1101/2020.12.09.20246520

**Authors:** Melissa Richard-Greenblatt, Matthew J. Ziegler, Valerie Bromberg, Elizabeth Huang, Hatem Abdallah, Pam Tolomeo, Ebbing Lautenbach, Laurel Glaser, Brendan J. Kelly

**Author notes:** **CORRESPONDING AUTHOR CONTACT** Brendan J. Kelly, MD, MS. **AUTHORS’ CONTRIBUTIONS** MRG - study design, data collection, manuscript. MJZ - study design, data analysis, manuscript. VB - data collection, manuscript. EH - data collection, manuscript. HA - data collection, manuscript. PT - data collection, manuscript. EL - data analysis; manuscript. LG - study design, manuscript. BJK - study design, data analysis, manuscript. **DISCLOSURES** The authors report no relevant disclosures. **DATA AVAILABILITY** Data, analysis scripts, and model code are available at github.com/bjklab.

## Abstract

**Background:** The SARS-CoV-2 reverse-transcription polymerase chain reaction (RT-PCR) cycle of threshold (Ct) has been used to estimate quantitative viral load, with the goal of targeting isolation precautions for individuals with COVID-19 and guiding public health interventions. However, variability in specimen quality can alter the Ct values obtained from SARS-CoV-2 clinical assays. We sought to define how variable nasopharyngeal (NP) swab quality impacts clinical SARS-CoV-2 test sensitivity.

**Methods:** We performed amplification of a human gene target (β-actin) in parallel with a clinical RT-PCR targeting the SARS-CoV-2 *ORF1ab* gene for 1311 NP specimens collected from patients with clinical concern for COVID-19. We evaluated the relationship between NP specimen quality, characterized by high Ct values for the human gene target β-actin Ct, and the probability of SARS-CoV-2 detection via logistic regression, as well as the linear relationship between SARS-CoV-2 and β-actin Ct.

**Results:** Low quality NP swabs are less likely to detect SARS-CoV-2 (odds ratio 0.654, 95%CI 0.523 to 0.802). We observed a positive linear relationship between SARS-CoV-2 and β-actin Ct values (slope 0.169, 95%CI 0.092 to 0.247). COVID-19 disease severity was not associated with β-actin Ct values.

**Conclusions:** Variability in NP specimen quality accounts for significant differences in the sensitivity of clinical SARS-CoV-2 assays. If unrecognized, low quality NP specimens, which are characterized by a low level of amplifiable human DNA target, may limit the application of SARS-CoV-2 Ct values to direct infection control and public health interventions.

## Introduction

As the COVID-19 pandemic continues to drive morbidity and mortality around the world, interest has grown in using SARS-CoV-2 reverse-transcription polymerase chain reaction (RT-PCR) cycle of threshold (Ct) values as a means of quantifying viral load (1, 2). It has been proposed that SARS-CoV-2 Ct values may correspond with viral burden and infectivity, and that SARS-CoV-2 values may be used to predict disease severity and guide isolation precautions for individuals with COVID-19 (3–7). SARS-CoV-2 Ct values have been shown to correspond with community COVID-19 burden, and it has also been proposed that community Ct values may help to guide non-pharmaceutical interventions to control COVID-19 (8).

We sought to understand the impact of nasopharyngeal (NP) specimen swab quality on the measurement of SARS-CoV-2 Ct and the sensitivity of virus detection. To collect an NP swab for SARS-CoV-2 testing, healthcare workers are instructed to advance a synthetic fiber swab with plastic or wire shaft through the nostril until contacting the posterior nasopharynx at a depth equal to the distance from the nostril to the opening of the ear, then to rub and roll the swab, leaving the swab in place for several seconds to collect secretions, before rotating the swab further as it is removed from the nostril (9). Variability in practice and patient tolerance of the procedure has been observed, and may impact the sensitivity of SARS-CoV-2 detection, as well as the cycle threshold (Ct) value observed when SARS-CoV-2 is detected (10–12).

To measure variability in the quality of NP swab collection, we performed amplification of a human gene target (β-actin) in parallel with RT-PCR targeting the SARS-CoV-2 *ORF1ab* gene. High β-actin Ct values have been previously validated as a marker of poor NP swab quality (2, 13). Below we report the relationship between quality of NP swab collection, sensitivity of SARS-CoV-2 detection, and the range of impact we expect sub-standard NP swab collection may exert on SARS-CoV-2 Ct values. We also examine the possibility of confounding by COVID-19 disease severity.

## Materials and Methods

### Study Design, Setting, and Population

We performed a retrospective cohort study, capturing consecutive SARS-CoV-2 RT-PCR tests performed at the Clinical Microbiology Laboratory of the Hospital of the University of Pennsylvania between March 26 and July 4, 2020. We included all SARS-CoV-2 RT-PCR results performed on NP specimens via the BD Max SARS-CoV-2 assay (Becton Dickinson) for which a positive PCR control analyte (MS2 phage DNA) was detected. A total of 1311 NP specimens were included. All specimens were assayed for β-actin and MS2 phage DNA in parallel with SARS-CoV-2. A waiver of informed consent was granted by the University of Pennsylvania Institutional Review Board (IRB protocols #843085 & 106 #843274).

### Causal Models

We hypothesized that β-actin and SARS-CoV-2 Ct values are related because poor NP specimen collection technique results in reduced capture of NP epithelial cells and SARS-CoV-2 alike. β-actin is a commonly used endogenous reference gene, used as an internal control for PCR reactions involving human specimens. This gene has been previously validated as a marker for the presence of nasal epithelial cells, and prior research has supported its use to assess the quality of self-collected midturbinate swabs (2, 13). We additionally considered the possibility of confounding by COVID-19 severity of illness. It is possible that those with more severe infection may have greater NP epithelial cell damage, resulting in greater detection of both PCR targets, irrespective of sampling technique.

### Clinical Data Collection

To evaluate the possibility of confounding by disease severity, we measured two independent markers of respiratory illness: (1) the minimum room-air oxygen saturation recorded within 2 days of SARS-CoV-2 testing, and (2) whether infiltrates were observed chest computerized tomography (CT) imaging performed within 7 days of SARS-CoV-2 testing. Per Centers for Disease Control and Prevention (CDC) guidelines(14), we considered room-air oxygen saturation < 94% indicative of severe respiratory illness. Radiology reports for CT imaging that described parenchymal lung disease, including “infiltrates”, “pneumonia”, “groundglass”, or other “opacities”, were considered indicative of severe respiratory illness. The presence of lung nodules, lung masses, chronic airway disease including bronchiectasis, emphysematous changes, or pleural effusions in the absence of parenchymal disease as described above, were not considered indicative of severe acute respiratory illness.

### Specimen Collection, Processing, and RT-PCR Assay

Specimens were collected during routine clinical practice using a nylon flocked mini-tip swab collected in VTM or saline. Healthcare providers obtained samples using CDC guidelines for NP samples collection. Samples were transported to the laboratory at ambient temperature and stored at 4°C if not run immediately. Exk TNA2 extraction reagent kits (Becton Dickinson) for the BD MAX open system reagent suite were used for the lab-developed SARS-CoV-2 assay based on a previously described assay (15, 16). The BD MAX system was set to run type 1 workflow. PCR conditions consisted of a reverse transcriptase step (600s at 58°C, 1 cycle), denaturation step (60s at 98°C, 1 cycle) and extension steps (10s at 98°C followed by 40s at 63°C, 40 cycles). Two different sets of primer/probe master mix were prepared and 12.5 µl was aliquoted into BD MAX 0.3 mL snap-in conical tubes for storage at −70°C prior to use. The LUNA Universal Probe One-Step RT-qPCR kit (New England Biolabs) was used to prepare the master mix according to manufacturer guidelines with modified primer and probe concentrations. Master mix 1 was composed of the SARS-CoV-2 orf1ab target (0.6 µM primers and 0.2 µM probe, sequences: unpublished data), and the internal processing control MS2 bacteriophage (0.1 µM primers and probe, sequences) (17). Master mix 2 contained the β-actin primers (0.6 µM) and probe (0.2 µM) (18). Samples were prepared by adding 200µl of NP specimen and 20 µl of specimen processing control (5×106 pfu/mL MS2 Phage; Zeptometrix) to an Exk TNA2 sample buffer tube. Sample buffer tubes containing patient specimens were loaded onto the BD MAX System racks along with the Exk TNA2 test strips. Master mix 1, neutralization buffer (25 µl NucliSENS easyMAG Extraction Buffer 3, Biomerieux) and master mix 2 were snapped in to open positions 2 to 4, respectively, on the test strip prior to loading the rack onto the BD MAX system. All NP samples for which the specimen processing control target (MS2) was detected were included in the study (n=1311). Ct values for all three targets (β-actin, MS2 and SARS-CoV-2) were recorded.

### Definition of Exposures and Outcomes

The primary exposure of interest was the β-actin Ct value, a surrogate for the quality of NP swab collection. The primary outcome of interest was SARS-CoV-2 Ct value.

### Statistical Methods

Data were organized using R statistical software version 3.6.1 (19), and plots generated using the “ggplot2” package (20). Where β-actin and SARS-CoV-2 were not detected, Ct values were imputed as 40 cycles. We examined (1) the linear relationship between β-actin and SARS-CoV-2 Ct values, as well as (2) the impact of β-actin Ct on SARS-CoV-2 detection using Bayesian linear and generalized-linear mixed effects models, which were fit using Stan Hamiltonian Monte Carlo (HMC) version 2.21, via the “brms” package with default weakly-informative priors (21, 22). Prior predictive modeling was performed, and models were fit with 4 chains of 1000 iterations, confirmed with HMC diagnostics (no divergent iterations, Rhat statistic < 1.1 for all parameters, and E-BFMI > 0.2) (23–25). We examined parameter distributions at 50%, 80%, and 95% posterior credible intervals to understand the relationship between exposure and outcome variables.

### Power and Sample Size

We estimated the necessary cohort size based on the anticipated effect of poor NP swab quantity (26). We anticipated that approximately 800 subjects would detect a 10% reduction in sensitivity of SARS-CoV-2 detection related to a β-actin Ct increase of 10, with credible intervals precision ensuring type S error < 5% (27, 28). We targeted enrollment of 10% more subjects to allow for a margin of error in that estimate, and we exceeded our enrollment target.

### Availability of Data

Data, analysis scripts, and model code are available at github.com/bjklab.

## Results

### SARS-CoV-2 Detection and Cycle of Threshold Range

Of 1311 tested specimens, 138 were found to have detectable SARS-CoV-2 within 40 cycles of PCR. Among these specimens, median SARS-CoV-2 Ct was 28.15 (IQR 20.5 to 32.98). No secular trend was observed between calendar time from local onset of COVID-19 cases and SARS-CoV-2 Ct values during the study period (Pearson correlation 0.18). **Figure 1** depicts the distribution of SARS-CoV-2 Ct values.

**Figure 1:**
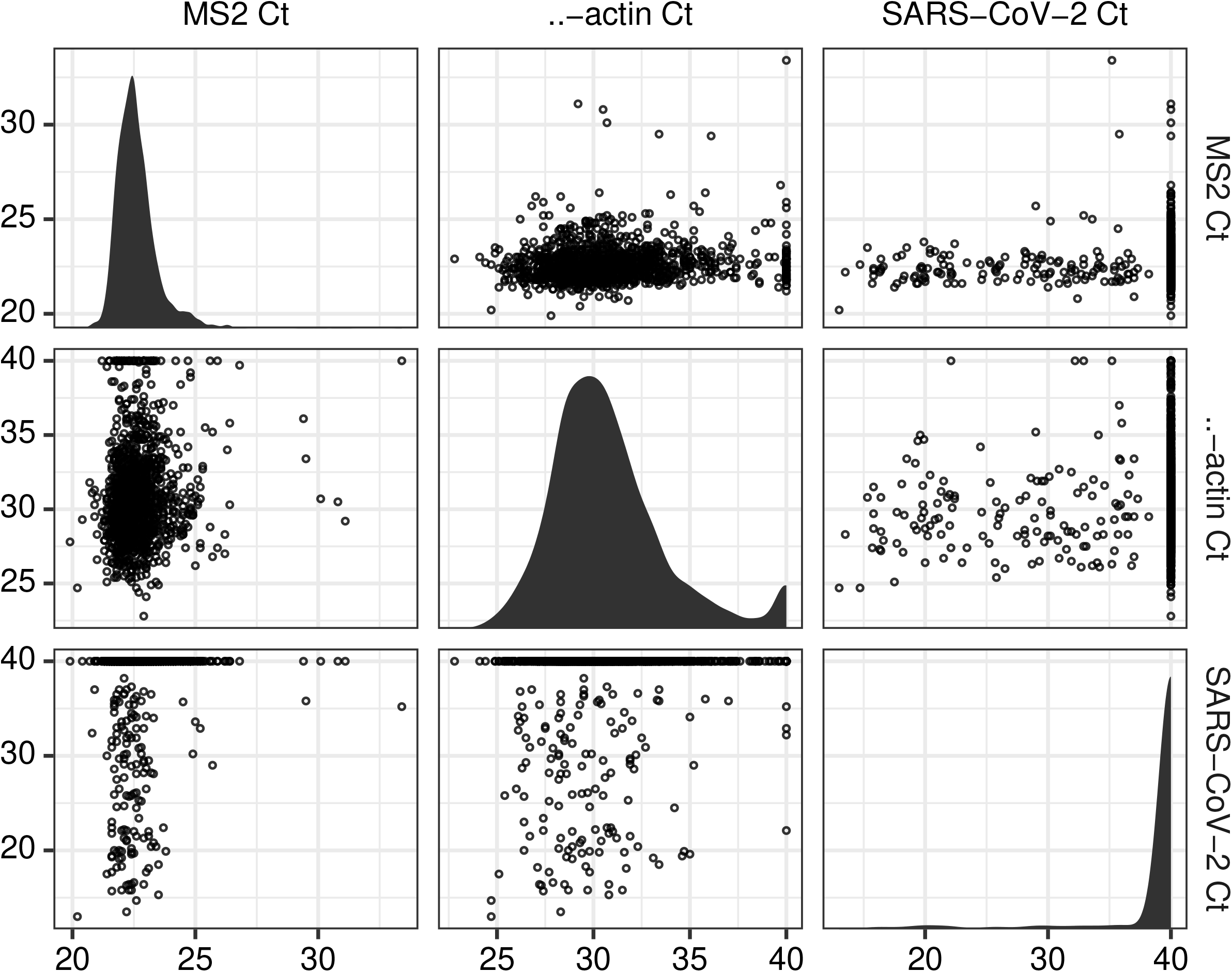
Distribution and relationships of Ct values for SARS-CoV-2, β-actin, and MS2 DNA positive control. A matrix plot depicting the observed cycle of threshold values for SARS-CoV-2 RT-PCR, with MS2 DNA positive control and β-actin specimen quality control over 1311 consecutive clinical assays run between March 26 and July 4, 2020. Panels on the diagonal present the distribution of each target’s Ct values. Panels off the diagonal present the relationship between Ct values for each pair of targets. Ct for specimens without detectable SARS-CoV-2 or β-actin were imputed at 40 cycles.

### Relationship between β-Actin and SARS-CoV-2 Cycle of Threshold

We evaluated the relationship between NP specimen quality, meaured by β-actin Ct value, and SARS-CoV-2 test sensitivity with logistic regression, and we found that increasing β-actin is significantly associated with reduced detection of SARS-CoV-2 (odds ratio 0.654, 95%CI 0.523 to 0.802). **Figure 2** depicts the relationship between β-actin and SARS-CoV-2 detection probability. We further evaluated the linear relationship between β-actin and SARS-CoV-2 Ct values with linear regression, and we found that SARS-CoV-2 Ct increases significantly with β-actin Ct (slope 0.169, 95%CI 0.092 to 0.247). A linear model restricted to include only the 134 specimens within which both SARS-CoV-2 and β-actin were detectable (i.e., Ct < 40) also found that SARS-CoV-2 Ct increased with β-actin, but this relationship did not have high posterior certainty (slope 0.254, 95%CI −0.23 to 0.73), and linear model fit was poor.

**Figure 2:**
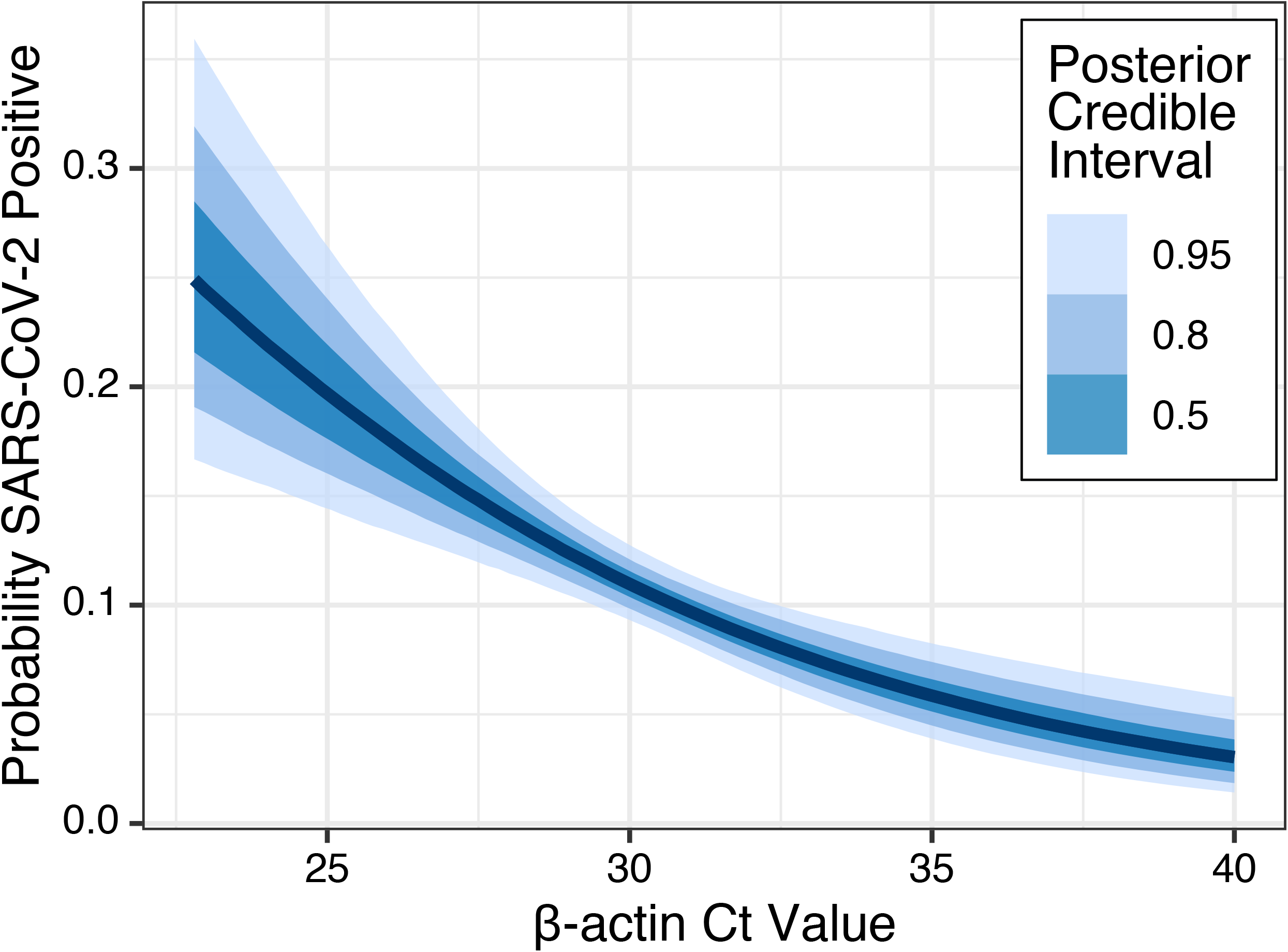
Relationship between β-actin and SARS-CoV-2 detection probability. Binomial logistic regression relating SARS-CoV-2 detection to β-actin Ct value reveals a negative association, with high β-actin Ct (i.e., low quality) NP specimens less likely to detect SARS-CoV-2. The absolute probabiity of SARS-CoV-2 detection is presented in relation to the observed range of β-actin Ct values.

### Impact of Poor NP Specimen Quality on SARS-CoV-2 Detection Sensitivity

To understand the potential impact of poor NP specimen quality, we evaluated the change in probability of SARS-CoV-2 detection as β-actin Ct increases. We found that a 4-Ct increase in β-actin, from Ct of 28 to Ct of 32 (roughly from the first quartile of observed β-actin Ct values to the third quartile) results in a 5.4% (95%CI 2.7% to 8.2%) decreased probability of SARS-CoV-2 detection.

### Impact of Disease Severity on Relationship Between β-Actin and SARS-CoV-2 Cycle of Threshold

Considering the possibility that the observed association between SARS-CoV-2 and β-actin Ct values is confounded by respiratory illness severity, we evaluated the relationship between β-actin Ct and independent markers of respiratory illness. Oxygen saturation data were available for 428 (32.6%) subjects; chest CT imaging was available for 111 (8.5%) subjects. Linear regression relating β-actin Ct values to oxygen saturation revealed no significant association, and the point estimate of association ran counter to concern for confounding by disease severity. Lower oxygen saturation was in fact associated with higher β-actin Ct values (less β-actin amplicon), with linear regression slope −0.04 (95%CI −0.273 to 0.182). Similarly, we found that the presence of parenchymal lung disease on chest CT radiography reports had no significant association with β-actin Ct values, and that the point estimate of association actually suggests lung parenchymal infiltrates are associated with higher β-actin Ct values (less β-actin Ct amplicon) with linear regression slope 0.428 (95%CI −0.655 to 1.61). These analyses of independent markers of severe respiratory disease suggest that it is NP specimen quality, not disease severity, that drives the association between SARS-CoV-2 and β-actin Ct values.

## Discussion

In this study, we found that higher β-actin Ct values, which have been previously validated as a marker of low NP swab quality (2, 13), were associated with reduced probability of SARS-CoV-2 detection (odds ratio 0.654, 95%CI 0.523 to 0.802) and with higher SARS-CoV-2 Ct values (linear regression slope 0.169, 95%CI 0.092 to 0.247). In our cohort, we observed 10.5% of tested NP specimens had detectable SARS-CoV-2. Based on the observed impact of NP specimen quality, we estimate that the reduction of quality NP specimen collection that results in β-actin Ct increasing from 28 to 32 (roughly 25% to 75% percentile) decreases the absolute probability of SARS-CoV-2 detection by 5.4% (95%CI 2.7% to 8.2%). This finding has several important implications. First, the correlation between β-actin Ct and SARS-CoV-2 suggests that quantitative interpretation of SARS-CoV-2 human specimens may be enhanced by adjusting for the β-actin Ct. Second, the data support the concern that poor specimen collection may contribute to false-negative results. The concern of false-negative NP SARS-CoV-2 testing has led to the recommendation to retest patients with moderate to high clinical suspicion of COVID-19 (29, 30). Reporting the β-actin Ct, or a β-actin-adjusted SARS-CoV-2 Ct may allow clinicians to better interpret specimen quality when considering retesting.

We considered the possibility that the observed relationship between SARS-CoV-2 and β-actin Ct values might be confounded by respiratory disease severity, but we found no significant association between independent markers of severe respiratory disease and lower β-actin Ct values. However, several limitations of our analysis must be acknowledged. Oxygen saturation data and chest CT radiography reports were only available for a small percentage (32.6% and 8.5%, respectively) of our subjects. Subject demographics and medical comorbidities could not be ascertained for subjects, so unmeasured confounders may contribute to the observed association.

Nevertheless, we believe that the observed association between NP specimen quality and SARS-CoV-2 RT-PCR sensitivity is an important finding. From 1311 NP specimens submitted for SARS-CoV-2 testing, we have quantified the variation in specimen quality measured by β-actin Ct value, and we have defined the impact of the observed variation on test sensitivity and SARS-CoV-2 Ct values.

SARS-CoV-2 Ct values have shown promise as a means to roughly quantify viral burden and so to guide infection control and public health interventions (1, 2, 4–8). However, variability in NP specimen collection may exert large effects on observed SARS-CoV-2 Ct values, limiting these useful applications. As testing efforts expand, infrastructure to ensure quality sample collection must expand as well (9). Concurrent measurement of a β-actin human gene target may provide a means to recognize and adjust for variability in NP specimen quality.

## Data Availability

Data, analysis scripts, and model code are available at https://github.com/bjklab/Impact-of-NP-Specimen-Quality-on-SARS-CoV-2-Test-Performance

## Acknowledgements

BJK, MJZ, and EL are supported in part by a CDC Cooperative Agreement FOA#CK16-004 “Epicenters for the Prevention of Healthcare Associated Infections”. BJK is supported by the National Institute for Allergy and Infectious Diseases (K23 AI121485 and L30 AI120149). MJZ is supported by the National Institute for Allergy and Infectious Diseases (K23 AI143925).

